# Rethinking anomia across the frontotemporal dementia spectrum: marker of language dysfunction or global cognitive decline?

**DOI:** 10.64898/2026.05.14.26353233

**Authors:** Shalom K. Henderson, Marissa Russell-Meill, Erie Shivers, Divya Sivakumar, Swathi Kiran

## Abstract

**Background:** Anomia is common in frontotemporal dementia (FTD), although its clinical prominence varies by subtype, with the most marked impairment typically observed in primary progressive aphasia (PPA). It remains unclear whether naming impairment reflects language-specific impairment or broader cognitive severity, and how it relates to other cognitive domains across FTD syndromes.

**Methods:** Fifteen healthy controls and twenty-two individuals across the FTD spectrum, including variant-specified and unclassifiable (NOS) presentations, completed two confrontation naming tasks (Boston Naming Test and Multilingual Naming Test) and a global cognitive screening measure (Montreal Cognitive Assessment, MoCA). Patient participants additionally completed a standardized language battery (Western Aphasia Battery – Revised) and a comprehensive neuropsychological assessment (Uniform Data Set). Naming performance was compared between groups and associations with language severity, global cognition, and domain-specific cognitive functions were examined using regression analyses.

**Results:** Naming was impaired in patients relative to healthy controls but did not differ between patient groups. Naming was strongly associated with language severity, but not global cognition. A significant group-by-MoCA interaction indicated that MoCA was positively associated with naming only in the unclassifiable group. In addition, naming was associated with episodic memory across both verbal and non-verbal domains.

**Conclusions:** Naming in FTD primarily reflects language severity rather than global cognitive impairment. A robust association between naming and episodic memory suggests potential contributions from semantic cognition, shared frontally mediated retrieval processes, or parallel cognitive decline. These findings support the use of naming as a marker of language dysfunction while highlighting its relevance to broader cognitive systems in FTD.

**What is already known on this topic:** Although not a core diagnostic feature of the non-fluent/agrammatic variant of primary progressive aphasia (nfvPPA) or the behavioral variant of frontotemporal dementia (bvFTD), naming impairment is frequently reported in these groups, sometimes early in the disease course. More broadly, language deficits across FTD syndromes are heterogeneous and have been linked to global cognitive severity.

**What this study adds:** This study provides a systematic evaluation of naming across FTD, PPA, and unclassifiable (i.e., FTD/PPA NOS) presentations using validated language and cognitive measures, which are rarely applied together. We provide new evidence that naming in FTD primarily reflects language severity rather than global cognitive decline. Despite prior studies reporting episodic memory deficits in FTD syndromes such as bvFTD, the link between memory and naming has not previously been established, and here we identify a novel association with not only verbal, but also non-verbal domains.

**How this study might affect research, practice or policy:** While anomia may represent a transdiagnostic feature across FTD syndromes, its severity may index the degree of language impairment, supporting the use of naming as a practical marker of language severity in clinical assessment. The observed association with episodic memory provides a framework for interpreting memory profiles in the context of shared underlying mechanisms with naming, with implications for both clinical evaluation and future research.

## Introduction

Frontotemporal dementia (FTD) encompasses a group of neurodegenerative syndromes that are classified according to their predominant clinical features, including behavioral variant of FTD (bvFTD) and primary progressive aphasia (PPA). While PPA is defined by progressive speech and/or language difficulties, with naming impairment as a core feature in semantic (svPPA) and logopenic (lvPPA) variants, bvFTD is characterized primarily by behavioral and executive dysfunction.^1-4^ However, increasing evidence suggests that these syndromes are not fully dissociable, and that cognitive and language deficits may overlap across the FTD spectrum.^5,6^ One such overlapping feature is anomia.^7-9^ In PPA, naming impairment is central to the syndrome and its underlying mechanisms are relatively well characterized. In svPPA/semantic dementia, anomia reflects degradation of semantic representations, whereas in lvPPA it is typically attributed to impaired lexical access.^10,11^ Although not a core diagnostic feature of the nonfluent/agrammatic variant (nfvPPA), naming deficits are often reported and may reflect a combination of motor speech deficits, impaired lexical access, and/or emerging semantic degradation.^12^ By contrast, naming impairment in bvFTD is less understood. Despite not being included in current diagnostic criteria, anomia is frequently observed, sometimes early in the disease course,^13,14^ and has been linked to semantic involvement rather than purely executive dysfunction.^6,7^ More broadly, language deficits in bvFTD are common and heterogeneous,^15,16^ with evidence suggesting an association with global cognitive severity.^7,17,18^

In clinical practice, overlap in anomia across FTD subtypes can lead to diagnostic ambiguity. When a patient presents with a prominent behavioral profile but also demonstrates impaired naming, it may be unclear whether this reflects (i) bvFTD with co-occurring aphasia, (ii) a language-led variant of FTD such as svPPA, (iii) semantic-behavioral variant of FTD,^19^ or (iv) a later stage of disease in which language has become secondarily affected.^20^ In many cases, this distinction cannot be determined retrospectively, underscoring the need for a clearer framework to interpret naming impairment across the FTD spectrum.

Whether naming deficits in bvFTD reflect the severity of underlying language impairment has yet to be established. Anomia is among the most common features of post-stroke and progressive aphasias,^21^ and performance on picture naming tests is often used as a proxy for overall severity of language impairment in the former group.^22^ Accordingly, confrontation naming tasks are widely used in clinical and research settings, including post-stroke aphasia prognosis,^23^ treatment outcome studies,^24^ and longitudinal tracking in PPA.^25,26^ However, their interpretability in bvFTD remains uncertain: whether naming deficits reflect a primary language impairment or instead index broader cognitive decline is not known.^7^ As comprehensive language assessments are time-intensive and not routinely administered in bvFTD, clinicians often rely on brief measures such as naming, increasing the importance of understanding what these measures capture across FTD syndromes.

The present study addresses two important gaps in the literature. First, the extent to which naming performance reflects language-specific impairment versus broader cognitive decline remains inconclusive in FTD. Second, existing studies have not systematically examined the relationship between naming and validated measures of language and cognition across FTD syndromes, limiting insight into whether these associations generalize across clinical variants. To this end, the present study addressed three aims: (1) to determine how naming performance differs across a transdiagnostic cohort comprising bvFTD, nfvPPA, lvPPA, and unclassifiable FTD/PPA, relative to healthy controls; (2) to examine whether naming performance reflects overall language severity and/or global cognitive status, as indexed by the Western Aphasia Battery – Aphasia Quotient and Montreal Cognitive Assessment; and (3) to evaluate the extent to which naming performance is influenced by cognitive domains beyond language, as captured by National Alzheimer’s Coordinating Center Uniform Data Set composite scores. This approach enabled us to inform the clinical interpretation of naming performance and its utility across FTD syndromes.

## Materials and methods

### Participants

Individuals with a clinical diagnosis of FTD and PPA, including both variant-specified and unclassifiable presentations, were recruited from specialist clinics, aphasia and other community groups, and the FTD Disorders Registry. The cohort comprised 22 patients: seven with bvFTD, five with nfvPPA, three with lvPPA, and seven classified as FTD or PPA not otherwise specified (NOS).^27^ The inclusion of the NOS group was deliberate, as such patients are frequently excluded from research despite clear clinical evidence of FTD. At the time of assessment, they did not meet criteria for a specific subtype, although disease evolution may permit later diagnostic clarification in some individuals. Fifteen healthy controls were recruited from the Boston University volunteer panel, as well as through word-of-mouth and community advertisements.

Thirty participants identified as ‘White,’ three as ‘Black or African American,’ two as ‘more than one race,’ one as ‘Asian,’ and one as ‘American Indian.’ All but one participant reported English as their native language: the remaining participant reported Gujarati as their native language. Written informed consent was obtained from all participants, including permission to access medical records for patient participants. Medical records encompassing clinical data and diagnostic work-up (including structural MRI, FDG-PET, amyloid PET imaging, cerebrospinal fluid analyses, and genetic testing, if available) were systematically reviewed to verify diagnostic classification in accordance with established consensus criteria for bvFTD and PPA.^1,2^ Diagnoses were established by each patient’s treating neurologist. Symptom duration was derived from patient and caregiver report and subsequently cross-validated against clinical documentation. Genetic testing results were available for 10 patient participants; among these, C9orf72 expansion was identified in three participants with bvFTD or NOS.

For analytic purposes, participants with nfvPPA and lvPPA were combined into a single PPA subgroup given the small sample size. Although these variants differed in their core clinical features, most notably greater repetition impairment in lvPPA and motor speech and syntactic impairment in nfvPPA, they exhibited comparable naming performance (nfvPPA: Boston Naming Test (BNT) mean = 24.6, Multilingual Naming Test (MINT) mean = 29.6; lvPPA: BNT mean = 26.3, MINT mean = 28.0). Biomarker evidence of Alzheimer disease (AD) pathology was available for all three lvPPA participants: only one participant had cerebrospinal fluid biomarkers indicative of AD.

### Assessments

All participants underwent a comprehensive research evaluation. Naming was assessed using the 30-item odd version of the Boston Naming Test (BNT) and the Multilingual Naming Test (MINT). Language function was indexed by the Western Aphasia Battery – Revised Aphasia Quotient (WAB-R AQ), and global cognition by the Montreal Cognitive Assessment (MoCA) and the National Alzheimer’s Coordinating Center Uniform Data Set (UDS v3.0).

Domain-specific cognitive performance was further characterized using UDS composite scores spanning memory, attention, executive, and visuospatial domains.

All patient participants completed the BNT, MINT, WAB-R AQ, and MoCA. The UDS battery was completed by all but one patient due to severely limited verbal output; the Trail Making Test (part of the UDS battery) was discontinued in two participants due to task difficulty. Healthy control participants completed a subset of assessments, including both naming measures and the MoCA.

The UDS battery has established normative data accounting for systematic effects of age, sex, and education, implemented in a publicly available normative calculator.^28^ Consistent with prior work,^29^ domain-specific composite scores were derived by converting individual test scores to *z*-scores and averaging across measures within each cognitive domain. Importantly, because the language composite score incorporates the MINT (our primary dependent measure), language domain scores were not calculated to avoid circularity. The UDS variable names are indicated in brackets below for reference:

1. Memory Composite Score: Immediate Craft Story Recall (paraphrase scoring), Delayed Craft Story Recall (paraphrase scoring), Total Score for Delayed Recall of Benson figure
2. Attention Composite Score: Trail Making Test Part A (Adjusted), Forward Number Span Test (# of correct trials)
3. Executive Function Composite Score: Trail Making Test Part B (Adjusted), Backward Number Span Test (# of correct trials)
4. Visuospatial Composite Score: Total Score for Copy of Benson Figure

### Statistical analysis

Demographic and clinical characteristics were compared across groups using Kruskal-Wallis tests for continuous variables and Fisher’s exact tests for categorical variables, with *post hoc* comparisons conducted using Dunn’s test with Bonferroni correction.

To examine naming performance, complementary analyses were used to assess overall effects of disease (i.e., patients versus controls) and diagnostic differentiation. Naming scores (BNT and MINT) were compared (i) between healthy controls and all patients combined using Wilcoxon rank-sum test and (ii) across groups using Kruskal-Wallis test with *post hoc* pairwise comparisons using Dunn’s test with Bonferroni correction. Nonparametric tests were selected over parametric alternatives (e.g., ANOVA) due to small and unequal group sizes and non-normal distributions.

To examine the relationship between naming performance and overall language and global cognitive function, two separate multiple linear regression models were conducted, with BNT and MINT scores entered as dependent variables in independent analyses. In each model, WAB-R AQ, MoCA, and diagnostic group were included as predictors, along with group x WAB-R AQ and group x MoCA interaction terms to assess group-specific effects. The bvFTD group was used as the reference group in both models. Significant interactions were probed via simple slopes using estimated marginal trends, with Bonferroni correction applied.

To examine associations between naming performance and domain-specific cognition across diagnostic groups, separate linear regression models were fitted for each cognitive domain (excluding language as aforementioned), with MINT *z*-scores as the dependent variable and UDS domain-specific composite *z*-scores (memory, attention, executive, and visuospatial) as predictors, including interaction terms between composite scores and diagnostic group, with bvFTD serving as the reference group. Separate models were used to limit model complexity given the sample size and to reduce potential multicollinearity among cognitive domain measures. When significant effects were observed at the composite level, follow-up exploratory linear regression analyses examined relationships at the level of individual task *z*-scores to differentiate verbal and non-verbal contributions. Tasks were classified as verbal or non-verbal based on their primary response modality (e.g., story recall and digit span as verbal; figure recall and trails as non-verbal). *Post hoc* analyses used emmeans (estimated marginal means and trends) with Bonferroni correction.

## Results

### Demographic and clinical details

Demographic and clinical details for each group are shown in Table 1. There were no significant differences between groups on age, gender, handedness, and education. Self-reported symptom duration differed between patient groups, with bvFTD patients reporting longer symptom duration that those with PPA (*p* = 0.05). MoCA *z*-scores also differed overall; however, *post hoc* comparisons showed only marginal differences between healthy controls and the PPA (*p* = 0.09) and NOS (*p* = 0.08) groups. Notably, no significant between-group differences were observed in WAB-R or UDS composite *z*-scores, confirming comparable language and cognitive severity across patient groups.

**Table 1.** Demographic and clinical data of the study cohort.

|  | **Healthy control** | **bvFTD** | **FTD/PPA NOS** | **lvPPA & nfvPPA** | **P\*** |
| --- | --- | --- | --- | --- | --- |
| **Demographics** |  |  |  |  |  |
| N | 15 | 7 | 7 | 8 | - |
| Age (years) | 69.3 (7.6) | 61.1 (9.2) | 70.3 (12.8) | 73.9 (6.4) | ns |
| Gender (male/female) | 10/5 | 5/2 | 3/4 | 7/1 | ns |
| Handedness (right/left) | 13/2 | 5/2 | 6/1 | 7/1 | ns |
| Education (years) | 16.9 (2.7) | 14.9 (2.0) | 17.0 (2.5) | 17.9 (2.5) | ns |
| Symptom duration (years) | - | 9.0 (4.3) | 6.6 (3.2) | 4.5 (1.3) | 0.05 |
| Mutation (yes/no/unknown) | - | 1/4/2 | 2/1/4 | 0/2/6 | - |
| **NACC Uniform Data Set (Z-scores)** |  |  |  |  |  |
| MoCA | -0.1 (1.1) | -2.0 (3.5) | -2.0 (2.1) | -2.3 (2.9) | 0.03 |
| Memory composite |  | -1.7 (2.0) | -0.8 (1.0) | 0.1 (1.9) | ns |
| Attention composite |  | -1.2 (1.4) | -1.4 (0.2) | -1.5 (0.8) | ns |
| Executive composite |  | -1.4 (1.2) | -1.4 (0.6) | -1.8 (0.5) | ns |
| Visuospatial composite |  | -1.2 (3.3) | -0.1 (1.1) | -0.1 (0.8) | ns |
| **Western Aphasia Battery – Revised (Raw scores)** |  |  |  |  |  |
| WAB-R AQ (/100) |  | 90.7 (14.8) | 90.6 (10.6) | 86.1 (14.2) | ns |
| WAB-R Spontaneous Speech (/20) |  | 18.0 (3.6) | 18.0 (2.8) | 16.0 (4.1) | ns |
| WAB-R Comprehension (/20) |  | 18.4 (2.8) | 19.7 (0.7) | 17.6 (4.8) | ns |
| WAB-R Repetition (/10) |  | 9.4 (0.8) | 9.1(1.1) | 8.3 (1.4) | ns |
| WAB-R Naming (/10) |  | 8.8 (1.8) | 8.4 (1.7) | 9.0 (1.0) | ns |
| WAB-R Reading (/20) |  | 18.3 (2.9) | 19.3 (0.7) | 18.5 (2.9) | ns |
| WAB-R Writing (/20) |  | 17.7 (4.4) | 18.0 (3.5) | 15.8 (7.1) | ns |

Note: Mean and standard deviations are displayed. \*P-value for group-difference by Kruskal-Wallis or Fisher’s exact tests. bvFTD, behavioral variant of frontotemporal dementia; lvPPA, logopenic variant of primary progressive aphasia; MoCA, Montreal Cognitive Assessment; NACC, National Alzheimer’s Coordinating Center; nfvPPA, non-fluent/agrammatic variant of primary progressive aphasia; NOS, not otherwise specified; ns, not significant; PPA, primary progressive aphasia; WAB-R, Western Aphasia Battery – Revised.

### Between-group differences in naming performance

As shown in Figure 1, when comparing healthy controls and all patients, naming performance differed between the two groups, with lower scores observed in patients on both the BNT (Wilcoxon rank-sum, *p* = 0.05) and MINT (Wilcoxon rank-sum, *p* = 0.03). Across the four groups, there was no overall difference in BNT scores (Kruskal-Wallis, *p* = 0.19). In contrast, MINT scores differed across groups (*p* = 0.05), with *post hoc* Dunn’s tests (Bonferroni-corrected) showing a significant difference between healthy controls and the NOS group (*p* = 0.04).

**Figure 1.**
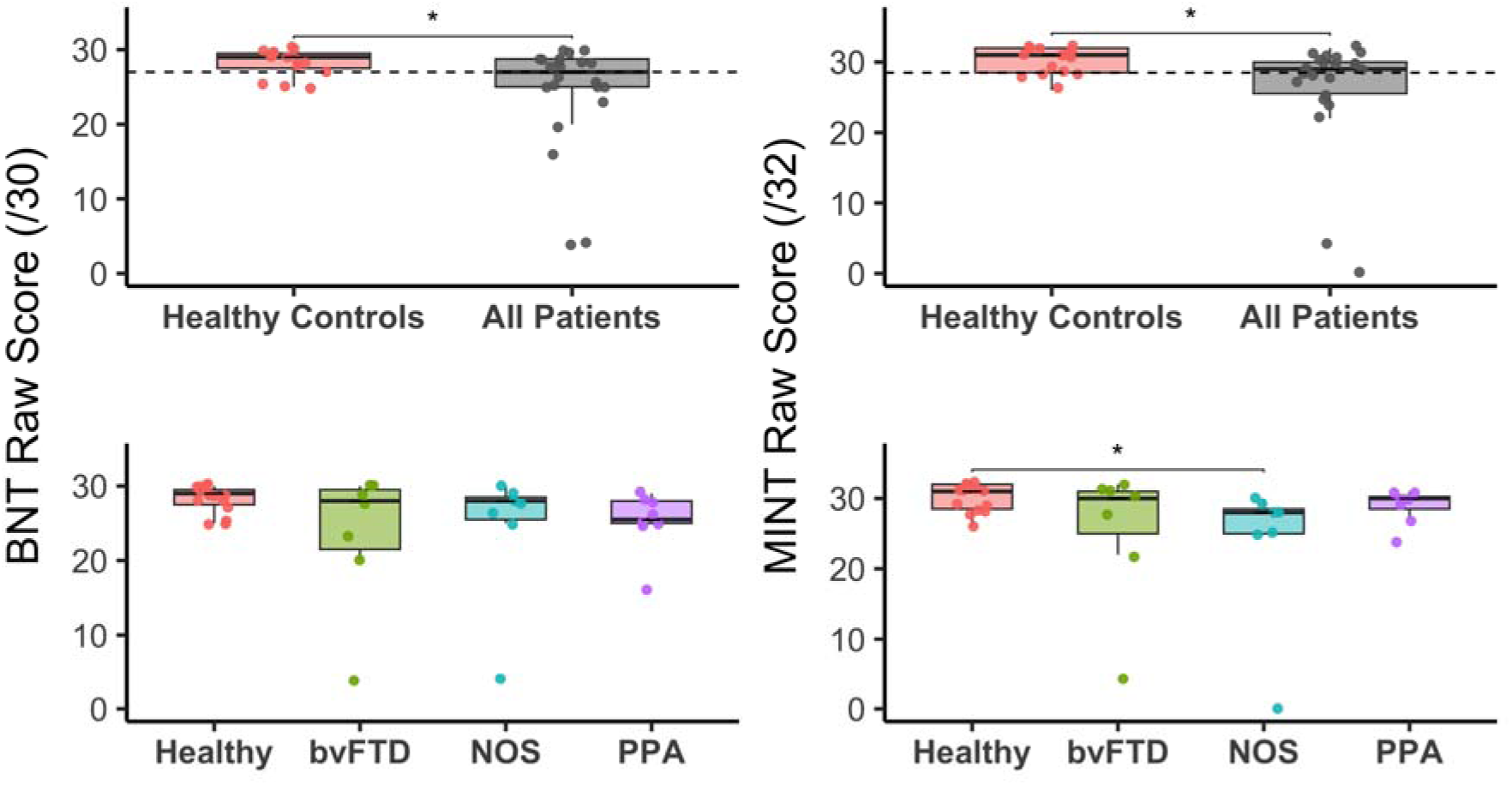
Naming performance on the Boston Naming Test (BNT) and Multilingual Naming Test (MINT) in healthy controls and patients. The top panels show comparisons between healthy controls and all patients combined: patients showed lower scores than healthy controls on both BNT (*p* = 0.05) and MINT (*p* = 0.03). The bottom panels show comparisons across groups: there was no difference in BNT (*p* = 0.19), but there was a significant difference for MINT (*p* = 0.05), with *post hoc* pairwise comparisons showing lower scores in the NOS group relative to healthy controls (*p* = 0.04). Dotted lines in the top panels indicate published normative cut-off scores.^30,31^

### Associations between naming, language severity, and global cognition

The model predicting BNT performance was significant (F(8,13) = 18.3, *p* < 0.001) and explained substantial variance (adjusted R² = 0.87). WAB-R AQ was a significant positive predictor (β = 0.54, *p* < 0.001), indicating that better language function was associated with higher BNT scores, whereas MoCA was not a significant predictor (*p* = 0.22). There was a significant group-by-MoCA interaction for the NOS group (*p* = 0.02). Simple slopes analyses showed that MoCA was positively associated with BNT performance in the NOS group (β = 1.12, 95% CI [0.45, 1.79]), but not in bvFTD (β = 0.21, 95% CI [-0.14, 0.57]) or PPA (β = 0.04, 95% CI [-0.45, 0.53]). *Post hoc* pairwise comparisons indicated that the MoCA and BNT association was stronger in NOS than in PPA (*p* = 0.05), with a trend toward a stronger association in NOS compared to bvFTD (*p* = 0.07).

Results for MINT were broadly similar. The model predicting MINT performance was significant (F(8,13) = 11.2, *p* < 0.001) and explained substantial variance (adjusted R² = 0.80). WAB-R AQ was again a significant positive predictor (β = 0.67, *p* < 0.001), whereas MoCA was not (*p* = 0.96). A significant group-by-MoCA interaction for the NOS group was observed (*p* = 0.02). Simple slopes analysis showed that MoCA was positively associated with MINT performance in the NOS group (β = 1.26, 95% CI [0.32, 2.19]), but not in bvFTD (β = −0.01, 95% CI [−0.51, 0.48]) or PPA (β = 0.43, 95% CI [−0.25, 1.11]), with a trend toward a stronger association in NOS compared to bvFTD (*p* = 0.07). In contrast to the BNT model, there was also a significant group-by-WAB-R AQ interaction for the PPA group (*p* = 0.002). Simple slopes analysis revealed that WAB-R AQ was positively associated with MINT performance in bvFTD (β = 0.67, 95% CI [0.38, 0.97]), but not in NOS (β = 0.20, 95% CI [−0.32, 0.73]) or PPA (β = −0.17, 95% CI [−0.52, 0.19]). *Post hoc* pairwise comparisons revealed that the WAB-R AQ and MINT association was significantly stronger in bvFTD than in PPA (*p* = 0.005).

Figure 2 shows the relationships between naming performance and WAB-R AQ (top panel) and MoCA (bottom panel). All points represent individual test scores. In the top panel, lines depict model-predicted values from the full regression model (BNT on the left; MINT on the right) as a function of WAB-R AQ, with MoCA held constant at the sample mean. In the bottom panel, lines depict model-predicted values as a function of MoCA, with WAB-R AQ held constant at the sample mean. Results from the simple slopes analyses are summarized in Figure 3.

**Figure 2.**
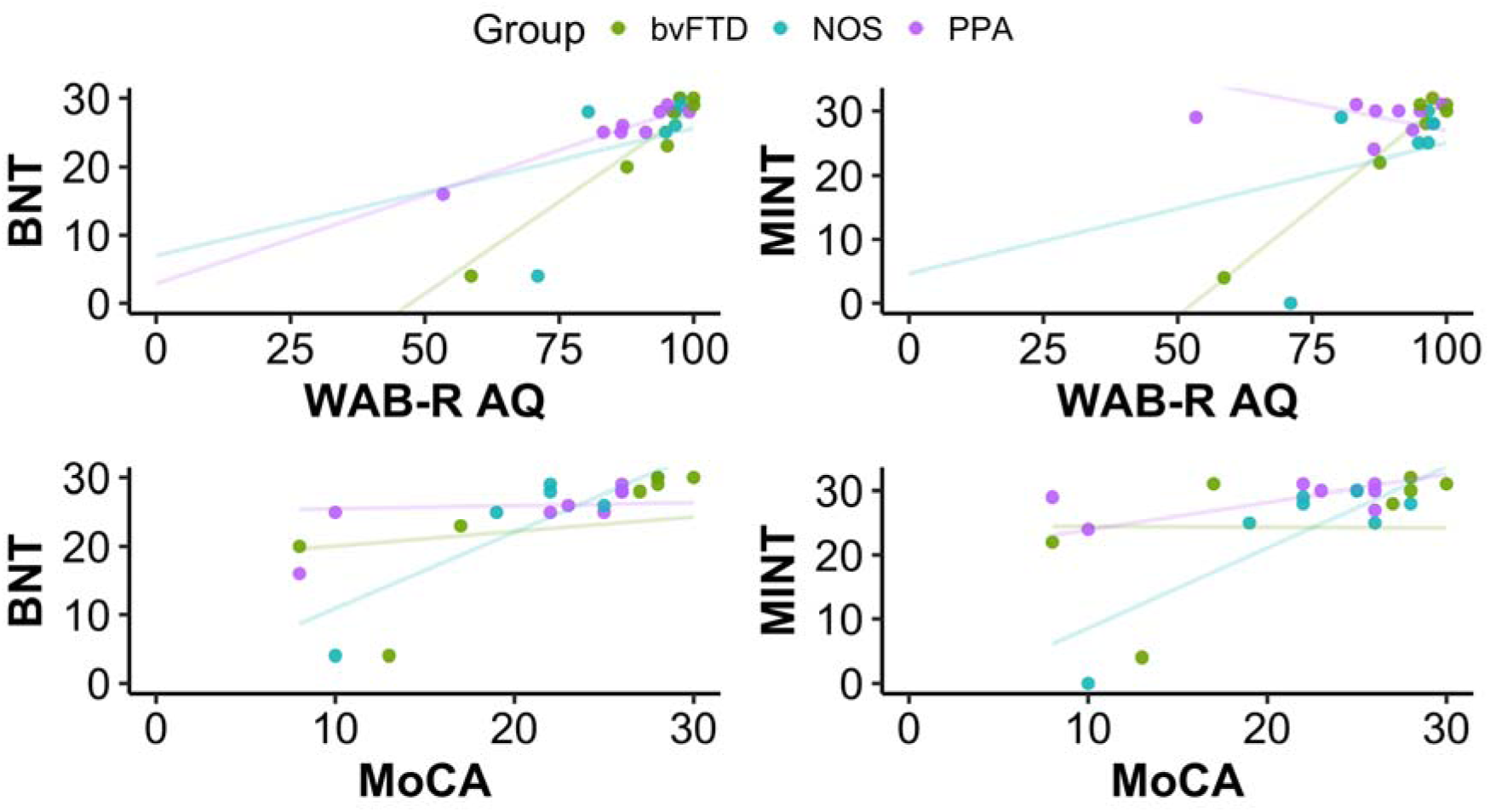
Associations between naming performance on the Boston Naming Test (BNT) and Multilingual Naming Test (MINT) and global language and cognitive measures. The top panels show associations with Western Aphasia Battery – Revised Aphasia Quotient (WAB-R AQ): lines represent model-predicted values from the full regression model with Montreal Cognitive Assessment (MoCA) held at the sample mean, indicating positive relationships between WAB-R AQ and both BNT and MINT. The bottom panels show associations with MoCA: lines represent model-predicted values with WAB-R AQ held at the sample mean; MoCA was not a significant predictor of naming performance. Points represent individual raw scores.

**Figure 3.**
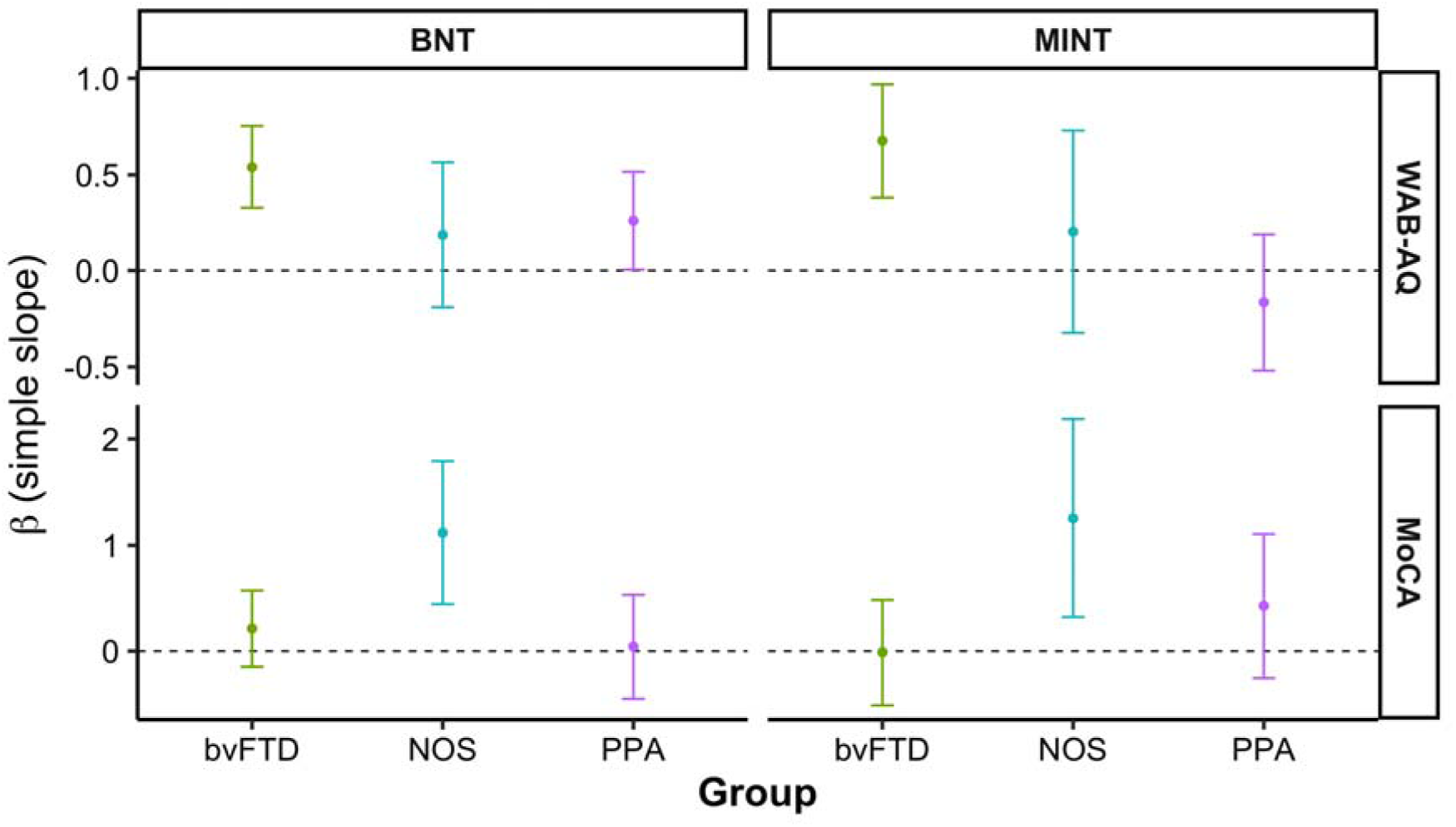
Simple slopes (β) for the associations between naming performance and global language and cognitive measures across diagnostic groups. The top panels show effects for Western Aphasia Battery – Revised Aphasia Quotient (WAB-R AQ): the bvFTD group show positive associations with both Boston Naming Test (BNT) and Multilingual Naming Test (MINT), NOS group shows weaker and more variable associations, and PPA group shows a positive association for BNT but a negative association for MINT. The bottom panels show effects for Montreal Cognitive Assessment (MoCA): NOS shows the strongest positive associations with both BNT and MINT, PPA shows modest positive associations, and bvFTD shows minimal or no association. Points indicate estimated slopes and error bars represent confidence intervals; the dashed line denotes zero.

### Contributions of non-language cognitive domains to naming performance

To examine associations between naming and domain-specific cognition across groups, separate linear regression models were run for each cognitive domain, with MINT *z*-scores as the dependent variable and UDS composite *z* -scores (memory, attention, executive, visuospatial) as predictors, including interactions with group. Only the memory composite model was significant (F(5,15) = 3.9, *p* = 0.01); models for attention, executive, and visuospatial domains were not significant. There were no significant interaction effects for the memory composite, indicating that higher memory performance was associated with better naming performance across groups.

Follow-up exploratory analyses examined individual memory tasks to distinguish verbal and non-verbal contributions. Figure 4 illustrates the relationships between MINT naming performance and memory tasks, including both composite and task-level *z*-scores. Immediate story recall (β = 1.43, *p* = 0.04), delayed story recall (β = 1.51, *p* = 0.05), and delayed figure recall (β = 1.52, *p* = 0.006) were all significant predictors of naming performance. A significant interaction was observed for delayed figure recall in the NOS group (*p* = 0.04); however, *post hoc* comparisons did not reveal significant group differences.

**Figure 4.**
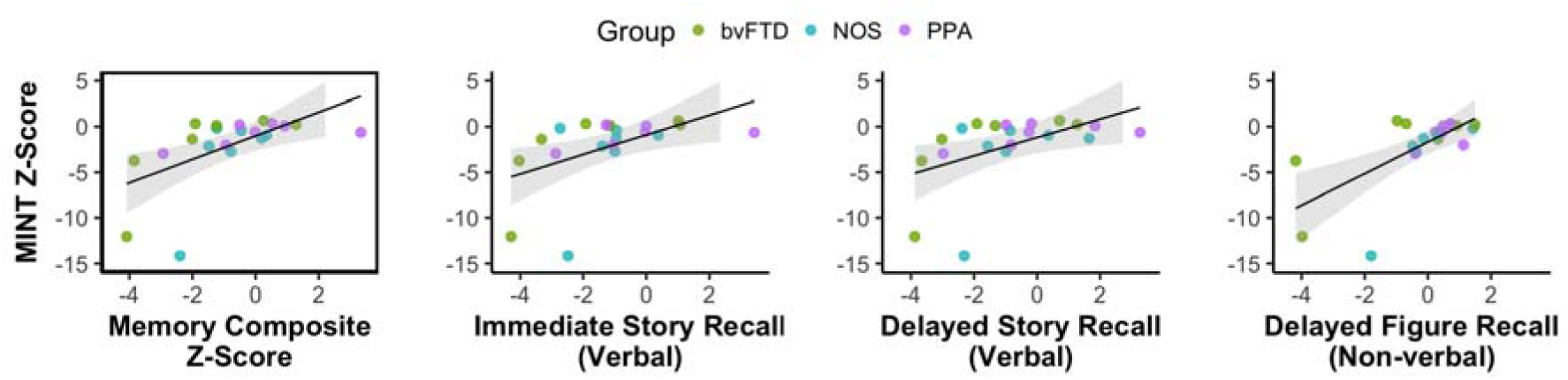
Association between Multilingual Naming Test (MINT) *z*-scores and Uniform Data Set (UDS) memory measures across the whole group. The boxed scatterplot on the left shows the relationship between MINT and the UDS memory composite *z*-scores. The remaining panels show associations with individual memory tasks: immediate story recall (verbal), delayed story recall (verbal), and delayed figure recall (non-verbal). Points represent individual *z*-scores, color-coded by diagnostic group, lines represent model-predicted values from linear regression models, and shaded areas indicate confidence intervals.

## Discussion

Anomia is prevalent across FTD syndromes,^9^ although its clinical prominence and relative severity varies by subtype, with the most marked impairment typically observed in svPPA/semantic dementia (SD) and less consistent deficits reported in nfvPPA and bvFTD. However, existing evidence is mixed, with some studies suggesting comparable rates of naming impairment in bvFTD.^7^ Critically, the extent to which naming performance reflects language-specific dysfunction versus broader cognitive decline in FTD remains unclear, and its relationship with validated measures of language and global cognition has rarely been examined within the same cohort. To this end, we examined naming performance in bvFTD, lvPPA, nfvPPA, and FTD/PPA NOS relative to age-matched healthy controls, and assessed its associations with language severity, global cognition, and non-linguistic cognitive domains. Three key findings are outlined below.

First, when comparing all patients to healthy controls, naming performance was impaired in patient groups for both BNT and MINT, supporting the clinical sensitivity of confrontation naming tasks to disease. However, naming did not differentiate between bvFTD, PPA, and unclassifiable cases. This finding should be interpreted with caution in light of the modest sample size and the absence of svPPA cases, in whom naming impairment is typically profound.^7,11^ Thus, inclusion of this group may have yielded greater diagnostic differentiation. Although group-level differences were not observed, naming performance showed considerable variability at the individual level as shown in Figure 1. While most participants were mildly anomic, more pronounced impairment was evident in a subset of individuals distributed across all diagnostic groups, rather than being confined to a specific group such as PPA. This pattern suggests that naming reflects a dimension of impairment that cuts across syndromes, prompting the question of whether it indexes language severity or broader cognitive decline, and whether these relationships vary by group.

Given the paucity of validated language assessments specifically sensitive to PPA and FTD, WAB-R remains a pragmatic index of overall language impairment despite recognized limitations.^32^ For both BNT and MINT, naming was strongly associated with WAB-R AQ even after controlling for MoCA. This finding extends observations from the post-stroke aphasia literature, where naming accuracy is well established as a proxy for the integrity of the language system and a robust indicator of overall aphasia severity.^23^ Our findings suggest that a similar relationship holds across the FTD spectrum, indicating that naming performance provides meaningful information about language severity in these disorders. Notably, the association between WAB-R AQ and naming performance differed by task in the PPA group. For MINT, this relationship was stronger in bvFTD than in PPA, whereas a comparable effect was not observed for the BNT. This pattern may suggest greater difficulty with the BNT in PPA; however, this interpretation warrants further investigation with larger samples. While MoCA was not associated with naming, we found an interaction in the NOS group, in whom naming performance showed a stronger association with global cognition. This may reflect more diffuse or mixed cognitive deficits, or less circumscribed language impairment, consistent with the absence of a clear syndromic classification.^33^ Alternatively, the NOS group may capture heterogeneous or transitional phenotypes, in which naming becomes more sensitive to broader cognitive decline.^27,34,35^

Our third and conceptually important finding was a robust association between naming and episodic memory. Across the cohort, MINT performance was strongly related to UDS memory composite scores, with consistent effects at the level of individual tasks, including verbal immediate and delayed recall and non-verbal delayed recall. Several mechanisms may account for this pattern. One possibility is a shared reliance on frontally mediated retrieval processes: both naming and episodic recall require strategic search, selection among competing representations, and retrieval under weak cue conditions, implicating prefrontal systems that are commonly affected in FTD.^36^ This interpretation aligns with prior work linking episodic memory deficits in bvFTD to disrupted attentional and executive control processes affecting strategic encoding and retrieval.^37^ However, the absence of an association between naming and executive composite scores suggests that this relationship is unlikely to reflect general executive dysfunction, and may instead reflect a more specific deficit in task-driven strategic search and retrieval processes.

An alternative account relates to impaired semantic cognition, which is common in FTD, as even ostensibly non-verbal memory tasks can draw on conceptual knowledge and semantic scaffolding strategies (e.g., drawing a big “box” with an “X” then a small “circle” on the right). Despite the absence of svPPA/SD in our sample, semantic degradation is well documented in bvFTD and may also be present in our NOS cases,^38^ potentially contributing to impairments in both naming and episodic memory. This account further highlights the importance of assessing naming in FTD, not only as measures of language function but also as potential proxies for broader semantic integrity.

Another potential explanation is that naming and episodic memory decline in parallel in FTD, such that anomic and amnestic features co-occur as part of a broader, multi-domain profile. Prior studies suggest that memory impairment in bvFTD may reflect disrupted hippocampal-based consolidation.^37,39^ However, in the absence of imaging and biomarker data, inferences should be made cautiously. With this in mind, the sample likely reflects predominantly FTLD-related rather than AD pathology: bvFTD is typically associated with tau, TDP-43, or FUS pathology, and although lvPPA is most strongly linked to AD, two of the three lvPPA participants were amyloid-negative. While the pathological substrate of NOS cases cannot be determined, the predominance of bvFTD and nfvPPA patients in the cohort suggests that AD pathology may be less prominent. In this context, if medial temporal structures are relatively preserved, naming and memory deficits may be better explained by retrieval-related or semantic processes than by primary encoding or storage impairment.

### Limitations

Several limitations should be considered. First, the sample size was modest, reflecting the inherent challenges of recruiting relatively rare conditions such as FTD and PPA. While typical for studies in this field, this may limit statistical power and the generalizability of findings. Second, the cohort did not include individuals with the svPPA/SD, which was not by design. Despite broad recruitment efforts across PPA subtypes, those who expressed interest were unable to complete the assessment due to disease severity and there were no other eligible participants. Given that svPPA/SD is classically and consistently associated with profound naming impairment, its absence limits the ability to fully characterize anomia across the FTD spectrum. Importantly, the comparative profile of naming performance in bvFTD relative to other PPA subtypes remains less well understood in the literature. In this context, the inclusion of bvFTD alongside nfvPPA and lvPPA represents a key strength of the present study. Third, the inclusion of an NOS group introduces diagnostic heterogeneity, as some individuals in the milder stages may evolve toward a more clearly defined clinical phenotype over time. However, rather than being a limitation, this reflects clinical reality: such cases are commonly encountered but often excluded from research despite clear evidence of FTD disease, and their inclusion enhances the ecological validity of the cohort and enables a more transdiagnostic examination of naming and cognition across the disease spectrum. Finally, three individuals in the sample had C9orf72 mutation. None were aphasic on the WAB-R, and all performed at or near ceiling on naming measures. Given the small number of cases, they are unlikely to have substantially influenced the overall pattern of results. Nonetheless, underlying genetic heterogeneity may contribute to variability in naming performance and its cognitive correlates and should be considered when interpreting these findings.^40^

## Conclusion

In FTD, naming is closely linked to language severity, independent of global cognitive status. The association between naming and episodic memory across verbal and non-verbal domains potentially reflects frontally mediated search and retrieval processes and/or semantic contributions. In clinical practice, these results provide a clearer framework for interpreting naming impairment, supporting its use as an index of language severity while recognizing its associations with broader cognitive processes such as episodic memory.

## Data Availability

The authors confirm that the data supporting the findings of this study are available within the article and its Supplementary material.

## Acknowledgements

We sincerely thank our participants and their families for supporting this work.

## Author Contributions

SKH contributed to the conceptualization, data curation, formal analysis, investigation, methodology, project administration, visualization, and writing of the original draft and review and editing of the final manuscript. MRM, ES, and DS contributed to the data curation and review of the text. SK contributed to funding acquisition, supervision, and review and editing of the text.

